# Effects of Right and Left Ventricular Pacing for Substrate Mapping Using Decrement-Evoked Potential Mapping in Patients with Scar-Related Ventricular Tachycardia

**DOI:** 10.1101/2025.08.09.25332991

**Authors:** Jae-Sun Uhm, Junbeom Park, Heeji Song, Jae-Kwang In, Junyoung Lee, Taehyun Hwang, Seunghoon Cho, Hanjin Park, Daehoon Kim, Hee Tae Yu, Tae-Hoon Kim, Chan Joo Lee, Jaewon Oh, Boyoung Joung, Hui-Nam Pak, Seok-Min Kang, Moon-Hyoung Lee

## Abstract

**Background:** The ventricular tachycardia (VT) substrate map is influenced by the rhythm during mapping. This study aimed to elucidate the effects of different pacing sites on substrate mapping using decrement-evoked potential (DEEP) mapping in patients with scar-related VT.

**Methods:** Patients with ischemic cardiomyopathy (ICM) or nonischemic cardiomyopathy (NICM) who underwent substrate mapping and ablation for scar-related VT were included. DEEP mapping was performed during right ventricular apex (RVA) and left ventricular outflow tract (LVOT) pacing. We analyzed the number, location, shape, and timing of lines of conduction block (LOB) using substrate maps obtained during RVA and LVOT pacing.

**Results:** A total of 19 patients (mean age, 62.7 ± 16.6 years; 17 males; 10 with ICM and 9 with NICM) were studied. DEEP mapping during RVA and LVOT pacing was performed in 16 patients. The number of pacemap-matching LOBs identified from the RVA S1, RVA S2, LVOT S1, and LVOT S2 maps were 0.61 ± 0.70, 1.24 ± 1.09, 1.00 ± 0.85, and 1.50 ± 1.17, respectively. The number of final pacemap-matching LOBs was 1.58 ± 1.07. Two LOBs were visible only during RVA pacing because they were parallel to the conduction direction. Six LOBs were visible only during LVOT pacing—five LOBs were parallel to the conduction direction, and one LOB was located at the wavefront collision area. During a mean follow-up of 7.6 ± 3.9, VT recurred in 26.3% of patients.

**Conclusion:** A high number of LOBs on critical substrates can be identified using two-site pacing DEEP mapping.

## INTRODUCTION

In patients with cardiomyopathy, the ventricular myocardium contains scar tissue which serves as a potential substrate for ventricular tachycardia (VT) reentry circuits. Activation and entrainment mapping are the most widely used techniques in scar-related VT. However, these techniques have limitations when mapping hemodynamically unstable, polymorphic, or pleomorphic VT.

With the development of high-resolution three-dimensional mapping technology, several mapping strategies, including isochronal late activation mapping (ILAM), decrement-evoked potential (DEEP) mapping, and magnetic resonance image (MRI)-based mapping, have been proposed for hemodynamically unstable scar-related VT. Nevertheless, optimal VT mapping and ablation strategies remain challenging and controversial.

The VT substrate map is influenced by the rhythm during mapping. However, the effects of pacing sites on DEEP mapping remain unknown. This study aimed to elucidate the effects of pacing sites on substrate mapping using DEEP mapping for scar-related VT in patients with ischemic cardiomyopathy (ICM) and nonischemic cardiomyopathy (NICM).

## METHODS

This cohort study was approved by the institutional review board (IRB number: 2025-1813-001) and conformed to the principles outlined in the Declaration of Helsinki. All patients provided written informed consent to undergo catheter ablation for VT.

Patients with ICM or NICM who underwent substrate mapping and catheter ablation for scar-related VT at two university hospitals between August 2024 and May 2025 were included. Patients aged ≤18 years and those who underwent catheter ablation for idiopathic VT, premature ventricular complexes, or nonsustained VT were excluded. Patients who underwent only VT activation mapping (without substrate mapping) were also excluded. Demographic and clinical data were collected from medical records.

For preprocedural risk assessment, the PAINESD score was used, with the following criteria: 5 points for pulmonary disease, 3 for age >60 years, 6 for ICM, 6 for New York Heart Association functional class ≥III, 3 for left ventricular ejection fraction (LVEF) <25%, 5 for VT storm, and 3 for diabetes mellitus.^4^ A PAINESD score ≥15 was considered high risk for poor outcomes. Figure 1 shows a flowchart of VT mapping and ablation in the present study. Procedures were performed under deep sedation. A His-RV catheter, a decapolar catheter with an inner lumen, and an octapolar microcatheter (Japan Lifeline, Tokyo, Japan) were placed in the right ventricular apex (RVA), coronary sinus, and anterior interventricular vein, respectively. Three-dimensional mapping was performed using the EnSite X system and an HD Grid mapping catheter (Abbott, Minneapolis, Minnesota, USA). The left ventricle (LV) was accessed via a transseptal or transaortic approach. A VT induction study was performed to confirm the QRS morphology of the clinical VT. If the VT was hemodynamically stable, activation mapping with entrainment mapping and catheter ablation of the critical isthmus were performed. If the VT was hemodynamically unstable or remained inducible after ablation for hemodynamically stable VT, substrate mapping was performed.

**Figure 1.**
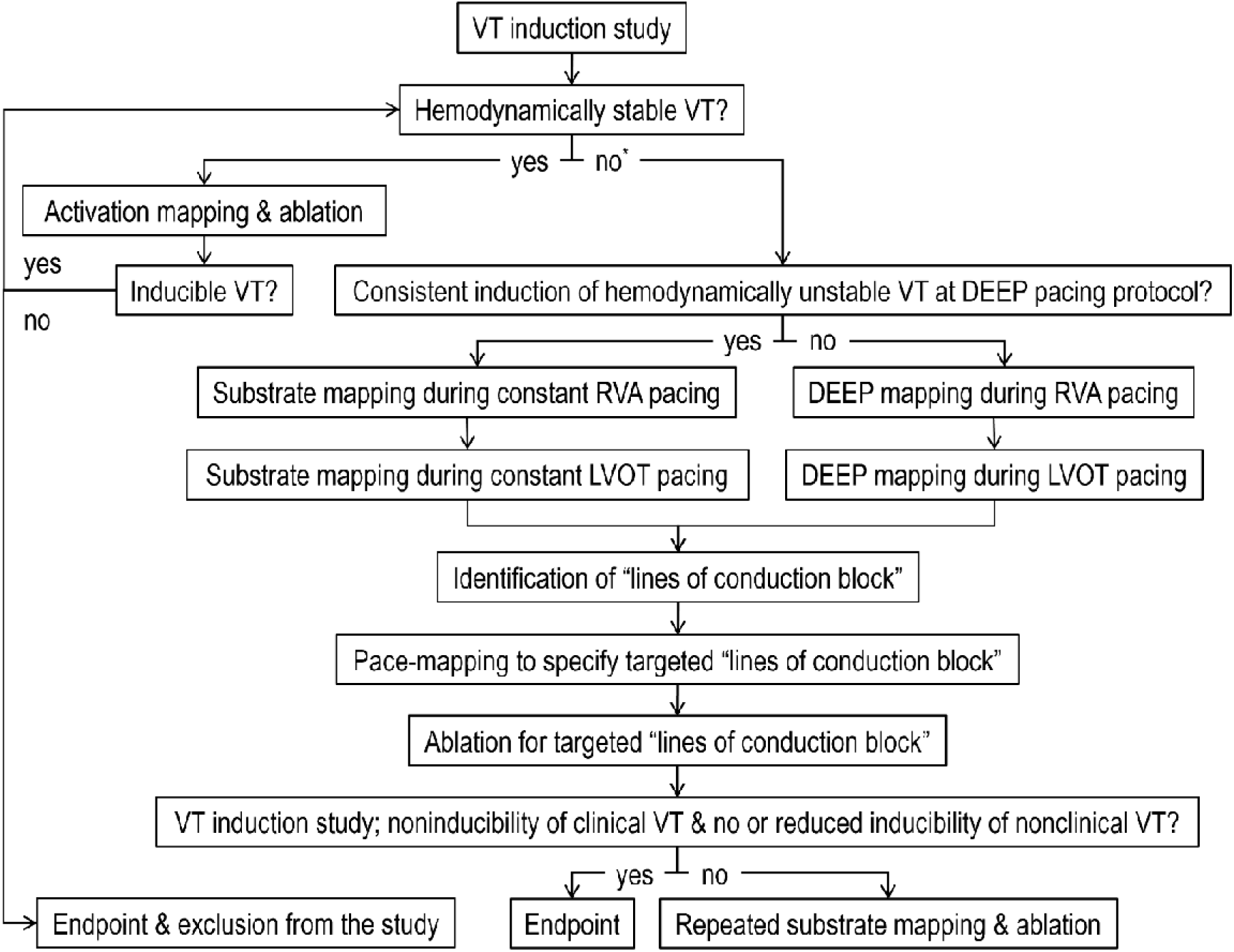
Flow chart of substrate mapping and ablation for scar-related ventricular tachycardia. DEEP, decrement-evoked potential; LVOT, left ventricular outflow tract, RVA, right ventricular apex; VT, ventricular tachycardia. *Hemodynamically unstable VT and no inducible VT.

DEEP mapping was conducted during RVA and left ventricular outflow tract (LVOT) pacing using the DEEP mapping pacing protocol (S1 = 500 ms; S2 = ventricular effective refractory period + 20 ms) with OT Near Field Technology (Abbott, St. Paul, Minnesota, USA). LVOT pacing was performed using the octapolar microcatheter placed in the anterior interventricular vein. If a hemodynamically unstable VT was consistently induced by the DEEP mapping pacing protocol, DEEP mapping was performed during RVA and LVOT pacing using a modified DEEP mapping pacing protocol (S1 = 500 ms, S2 = VT-inducing S2 interval + 20 ms). If hemodynamically unstable VT was still consistently induced by the modified DEEP mapping pacing protocol, VT substrate mapping was performed during RVA and LVOT pacing at constant intervals of 500 ms.

In bipolar voltage maps, scar and border zones were defined as voltages <0.5 mV and 0.5–1.5 mV, respectively.^5^ In unipolar voltage maps, the scar zone was defined as voltage <8.3 mV.^6^ Lines of conduction block (LOB), with or without a rotational activation pattern, were identified from the four substrate maps (RVA S1, RVA S2, LVOT S1, and LVOT S2 maps). Pacemapping using VT tachycardia cycle length was performed to identify targeted LOBs. Catheter ablation was conducted for LOBs that exhibited ≥90% pacemap matching with clinical VT.^7^

Radiofrequency energy was delivered using an ablation catheter (TactiCath, Abbott, Minneapolis, Minnesota, USA) with half-normal saline irrigation at 40–50 Watts for 90–120 s per application. A VT induction study was performed after catheter ablation. If VT was inducible, substrate mapping was repeated. The procedural endpoint was defined as non-inducibility of clinical VT and either non-inducibility or reduced inducibility of nonclinical VT. Reduced inducibility was defined as the absence of VT induction by extrastimuli that had previously induced VT before catheter ablation or VT induction by shorter or a greater number of extrastimuli.

After catheter ablation, amiodarone or mexiletine was continued for 3 months. If VT did not recur during this period, doses of amiodarone and mexiletine were reduced or discontinued. Medications for heart failure management (e.g., β-blockers) were continued. Patients were followed up at outpatient clinics at 3– to 6-month intervals. Implantable cardioverter defibrillators (ICDs) and cardiac resynchronization therapy defibrillators (CRT-D) were interrogated at every outpatient visit. VT recurrence was defined as ECG documentation of VT or appropriate ICD (or CRT-D) therapy for VT or ventricular fibrillation (VF).

We analyzed the number, location, shape, and activation timing of LOBs revealed by the RVA S1, RVA S2, LVOT S1, and LVOT S2 maps per patient. We also analyzed the number, location, shape, and activation timing of LOBs that were visible during only one pacing rhythm, along with the reasons for their invisibility during other pacing rhythms. The data were also analyzed and compared between patients with ICM and those with NICM.

### Statistical Analysis

Continuous variables are expressed as mean ± standard deviation and compared using the Mann–Whitney *U* test. Categorical variables are expressed as numbers (percentages) and compared using Fisher’s exact test. Statistical significance was set at P <0.05. Data were analyzed using the Statistical Package for the Social Sciences, version 27.0 (IBM Corp. Armonk, New York, USA).

## RESULTS

A total of 19 patients (mean age, 62.7 ± 16.6 years; 17 males) were included. The number of patients with ICM and NICM was 10 and 9, respectively. There were no significant differences in baseline characteristics between the ICM and NICM groups, except for the PAINESD score (Table 1). Substrate mapping and ablation outcomes are presented in Table 2. In seven patients, hemodynamically stable VT was induced, and VT activation mapping was performed. DEEP mapping during RVA and LVOT pacing was performed in 16 patients. In three patients, substrate mapping during S1 pacing with a constant interval was performed because hemodynamically unstable VT was consistently induced by the DEEP mapping pacing protocol.

**Table 1.** Baselines Characteristics.

|  | **Total** | **ICM** | **NICM** | **p-value** |
| --- | --- | --- | --- | --- |
|  | **(n = 19)** | **(n = 10)** | **(n = 9)** |  |
| **Age (years)** | 62.7 ± 16.6 | 69.3 ± 9.5 | 55.3 ± 20.0 | 0.327 |
| **Male, n (%)** | 17 (89.5) | 10 (100) | 7 (77.8) | 0.211 |
| **Etiology of cardiomyopathy** |  |  |  |  |
| Ischemic, n (%) | 10 (52.6) | 10 (100) | 0 (0) | <0.001 |
| Nonischemic, n (%) | 9 (47.4) | 0 (0) | 9 (100) | <0.001 |
| **Comorbidity** |  |  |  |  |
| Hypertension, n (%) | 2 (10.5) | 2 (20.0) | 0 (0.0) | 0.474 |
| Diabetes mellitus, n (%) | 6 (31.6) | 3 (30.0) | 3 (33.3) | >0.999 |
| Chronic kidney disease, n (%) | 4 (21.1) | 4 (40.0) | 0 (0.0) | 0.087 |
| Atrial fibrillation, n (%) | 4 (21.1) | 1 (10.0) | 3 (33.3) | 0.303 |
| **NYHA functional class** | 2.2 ± 0.8 | 2.3 ± 0.7 | 2.0 ± 0.9 | 0.430 |
| **LVEF (%)** | 34.2 ± 12.7 | 32.0 ± 13.4 | 36.6 ± 12.1 | 0.327 |
| **Electrical storm, n (%)** | 8 (42.1) | 4 (40.0) | 4 (44.4) | >0.999 |
| **PAINESD score** | 10.8 ± 6.6 | 14.3 ± 4.7 | 6.9 ± 6.4 | 0.033 |
| **Medication** |  |  |  |  |
| Amiodarone, n (%) | 19 (100.0) | 10 (100.0) | 9 (100.0) | 1.000 |
| Mexiletine, n (%) | 2 (10.5) | 1 (10.0) | 1 (11.1) | >0.999 |
| β-blockers, n (%) | 19 (100.0) | 10 (100.0) | 9 (100.0) | 1.000 |
| **CIED, n (%)** | 19 (100.0) | 10 (100.0) | 9 (100.0) | 1.000 |
| ICD, n (%) | 17 (89.5) | 9 (90.0) | 8 (88.9) | >0.999 |
| CRT-D, n (%) | 2 (10.5) | 1 (10.0) | 1 (11.1) | >0.999 |
| **Redo VT ablation, n (%)** | 8 (42.1) | 6 (60.0) | 2 (22.2) | 0.170 |
| **Follow-up duration (months)** | 7.6 ± 3.9 | 7.0 ± 4.3 | 8.3 ± 3.7 | 0.414 |

**Abbreviations:** CIED, cardiac implantable electronic device; CRT-D, cardiac resynchronization therapy-defibrillator; ICD, implantable cardioverter-defibrillator; ICM, ischemic cardiomyopathy; LVEF, left ventricular ejection fraction; NICM, nonischemic cardiomyopathy; NYHA, New York Heart Association; PAINESD, pulmonary disease, age, ischemic heart disease, NYHA, ejection fraction, VT storm, and diabetes mellitus; VT, ventricular tachycardia.

**Table 2.** VT Substate Mapping and Ablation Outcomes.

|  | **Total** | **ICM** | **NICM** |
| --- | --- | --- | --- |
|  | **(n = 19)** | **(n = 10)** | **(n = 9)** |
| **VT activation mapping, n (%)** | 7 (36.8) | 6 (60.0) | 1 (11.1) |
| **DEEP mapping during RVA & LVOT pacing, n (%)** | 16 (84.2) | 9 (90.0) | 7 (77.8) |
| **Epicardial mapping, n (%)** | 4 (21.1) | 0 (0.0) | 4 (44.4) |
| **Number of pacemap-matching LOBs** |  |  |  |
| RVA S1 | 0.61 ± 0.70 | 0.78 ± 0.83 | 0.44 ± 0.53 |
| RVA S2 | 1.24 ± 1.09 | 1.67 ± 1.12 | 0.75 ± 0.89 |
| LVOT S1 | 1.00 ± 0.85 | 1.29 ± 0.95 | 0.60 ± 0.55 |
| LVOT S2 | 1.50 ± 1.17 | 1.86 ± 1.07 | 1.00 ± 1.23 |
| Final LOBs | 1.58 ± 1.07 | 1.90 ± 1.20 | 1.22 ± 0.83 |
| LOBs additionally revealed by RVA S2 vs. RVA S1 | 0.69 ± 0.95 | 1.00 ± 1.20 | 0.38 ± 0.52 |
| LOBs additionally revealed by LVOT S1 vs. RVA S1 | 0.45 ± 1.04 | 0.67 ± 1.21 | 0.20 ± 0.84 |
| LOBs additionally revealed by LVOT S2 vs. RVA S1 | 1.00 ± 1.13 | 1.29 ± 1.11 | 0.60 ± 1.14 |
| LOBs additionally revealed by LVOT S2 vs. LVOT S1 | 0.50 ± 0.80 | 0.57 ± 0.79 | 0.40 ± 0.89 |
| **Endpoint achieved, n (%)** | 15 (78.9) | 8 (80.0) | 7 (77.8) |
| Noninducibility of any VT, n (%) | 8 (42.1) | 4 (40.0) | 4 (44.4) |
| Reduced inducibility of nonclinical VT, n (%) | 7 (36.8) | 4 (40.0) | 3 (33.3) |
| **Complications, n (%)** | **1 (5.3)** | **0 (0.0)** | **1 (11.1)** |
| Vascular, n (%) | 0 (0.0) | 0 (0.0) | 0 (0.0) |
| Cardiac tamponade, n (%) | 1 (5.3) | 0 (0.0) | 1 (11.1) |
| HF aggravation, n (%) | 0 (0.0) | 0 (0.0) | 0 (0.0) |
| **VT recurrence, n (%)** | **5 (26.3)** | **3 (30.0)** | **2 (22.2)** |

**Abbreviations:** DEEP, decrement-evoked potential; HF, heart failure; ICM, ischemic cardiomyopathy; LOB, line of conduction block; LVOT, left ventricular outflow tract; NICM, nonischemic cardiomyopathy; RVA, right ventricular apex; VT, ventricular tachycardia.

The numbers of pacemap-matching LOBs identified from the RVA S1, RVA S2, LVOT S1, and LVOT S2 maps were 0.61 ± 0.70, 1.24 ± 1.09, 1.00 ± 0.85, and 1.50 ± 1.17, respectively. The number of final LOBs identified from four substrate maps was 1.58 ± 1.07 overall, 1.90 ± 1.20 in patients with ICM, and 1.22 ± 0.83, in patients with NICM. Compared with RVA S1 map, the additional LOBs were 0.69 ± 0.95 from RVA S2, 0.45 ± 1.04 from LVOT S1, and 1.00 ± 1.13 from LVOT S2. The LVOT S2 map revealed the greatest number of LOBs with pacemap matching compared with the RVA S1, RVA S2, and LVOT S1 maps.

Table 3 lists the characteristics of LOBs with pacemap matching. Thirty LOBs were identified using substrate mapping. The activation timing of LOBs in the whole LV activation 34.6 ± 14.6% during RVA pacing and 28.4 ± 16.9% during LVOT pacing. The activation timing of all LOBs was within the first 65% of the entire LV activation. The frequencies of U-shaped, linear, and corridor-shaped (two parallel lines) LOBs were 56.7%, 36.7%, and 6.7%, respectively. All linear LOBs were oriented perpendicular or oblique to the direction of conduction. Across all patients, 66.7% of the LOB were located in scar zones and 33.3% in border zones. Compared with S1 mapping, S2 mapping revealed 14 additional LOBs (46.7%), deceleration augmentation in 6 LOBs (20.0%), and LOB elongation in 5 LOBs (16.7%).

**Table 3.**
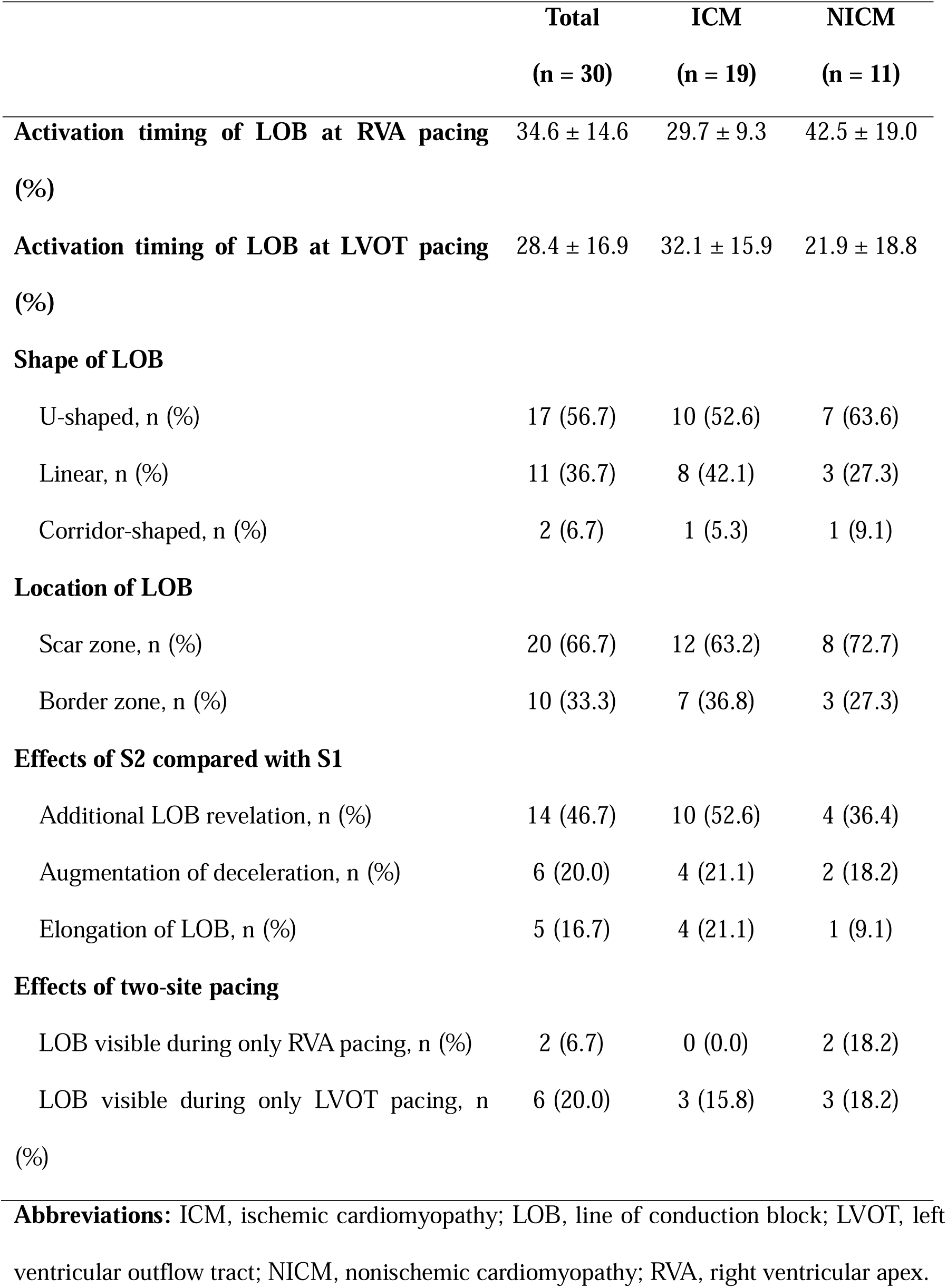
Characteristics of Lines of Conduction Block with Pacemap Matching.

Table 4 shows the number of LOBs that were visible during only one pacing rhythm and the reasons for their invisibility during other pacing rhythms. Two LOBs were visible during RVA pacing but not during LVOT pacing because they were parallel to the conduction direction. Six LOBs were visible during LVOT pacing but not during RVA pacing; five were parallel to the conduction direction, and one LOB was located in the wavefront collision area. The LOB at the wavefront collision area exhibited activation timing later than 65% of the whole LV activation time. Figures 2, 3, 4, 5, and 6 show the usefulness of LVOT S2, S2, LVOT, RVA, and LVOT pacing, respectively.

**Figure 2.**
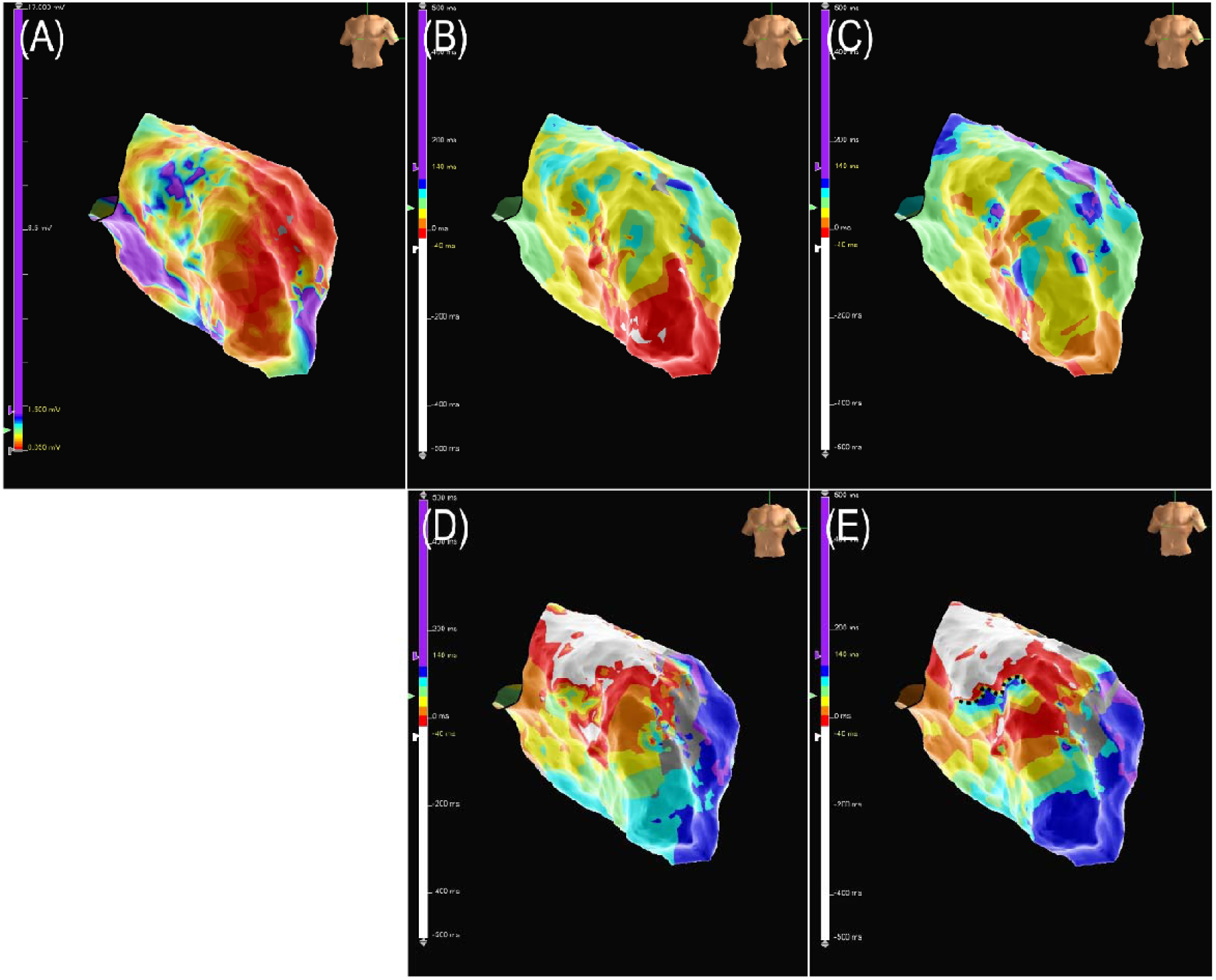
A male in his 60s with ischemic cardiomyopathy. (A) Voltage map; (B) RVA S1 map; (C) RVA S2 map; (D) LVOT S1 map; (E) LVOT S2 map. A LOB (black dotted line) at the LV mid-anterior wall was identified in the LVOT S2 map but not in the RVA S1, RVA S2, and LVOT S1 maps. In the RVA S1 and S2 maps, the LOB was not visualized because it was parallel to the conduction direction. Augmentation of deceleration at the LOB was observed in the LVOT S2 map compared with the LVOT S1 map. **Abbreviations:** LOB, line of conduction block; LV, left ventricle; LVOT, left ventricular outflow tract; RVA, right ventricular apex.

**Figure 3.**
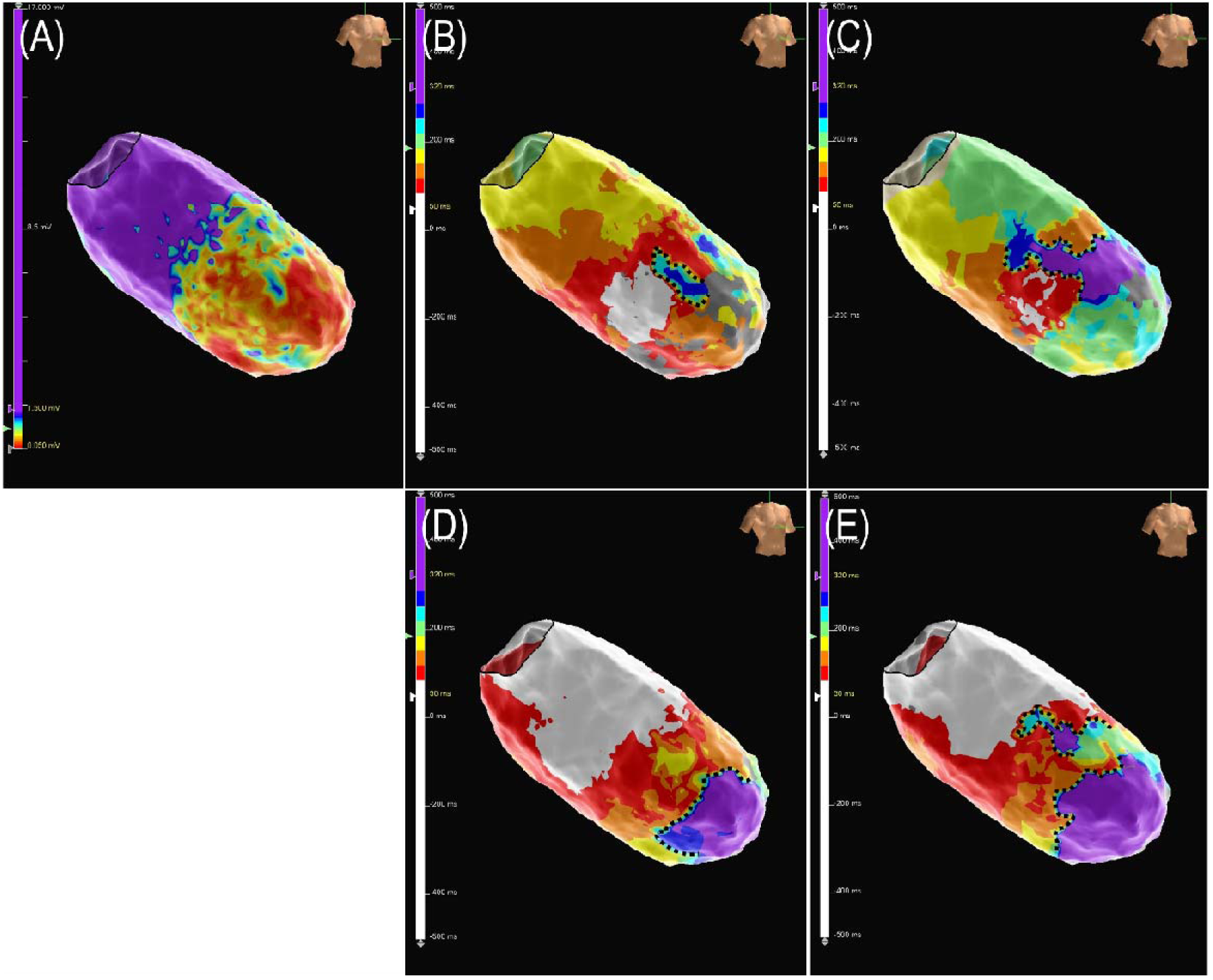
A male in his 70s with ischemic cardiomyopathy. (A) Voltage map; (B) RVA S1 map; (C) RVA S2 map; (D) LVOT S1 map; (E) LVOT S2 map. A corridor-shaped LOB (black dotted line) at the LV mid-anterior wall was identified in the RVA S1, RVA S2, and LVOT S2 maps, but not in the LVOT S1 map. A linear LOB (black dotted line) at the LV apical wall was identified in the LVOT S1 and LVOT S2 maps, but not in the RVA S1 and RVA S2 maps. These LOBs were not visualized because they were parallel to the conduction direction. Augmentation of deceleration at the LOBs was observed in S2 maps, compared with S1 maps. **Abbreviations:** LOB, line of conduction block; LV, left ventricle; LVOT, left ventricular outflow tract; RVA, right ventricular apex.

**Figure 4.**
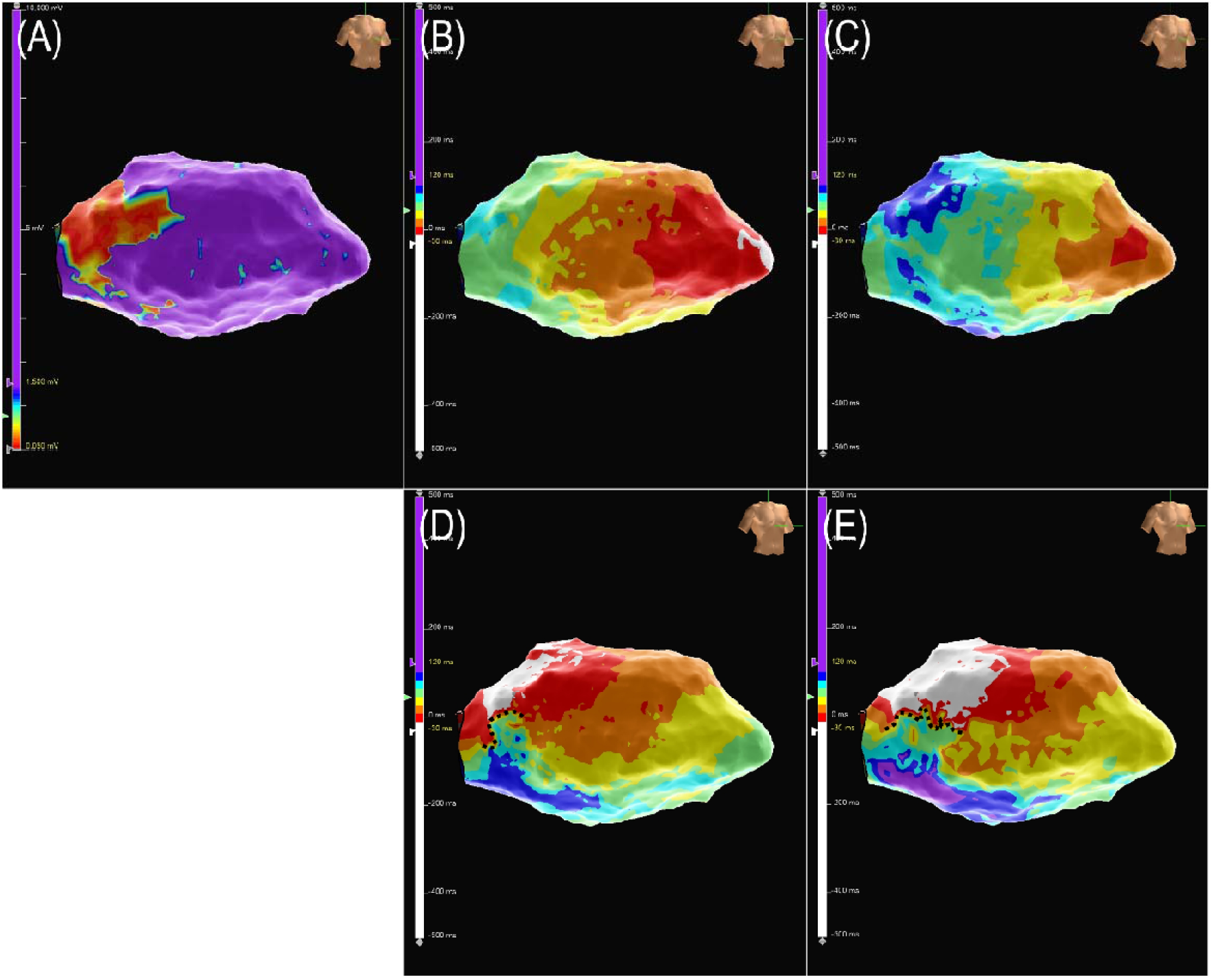
A male in his 70s with nonischemic cardiomyopathy. (A) Voltage map; (B) RVA S1 map; (C) RVA S2 map; (D) LVOT S1 map; (E) LVOT S2 map. A LOB (black dotted line) at the LV basal septal wall was identified in the LVOT S1 and LVOT S2 maps but not in the RVA S1 and RVA S2 maps. The LOB was not visualized during RVA pacing because it was parallel to the conduction direction. The LOB was elongated in the LVOT S2 map compared with the LVOT S1 map. **Abbreviations:** LOB, line of conduction block; LV, left ventricular; LVOT, left ventricular outflow tract; RVA, right ventricular apex.

**Figure 5.**
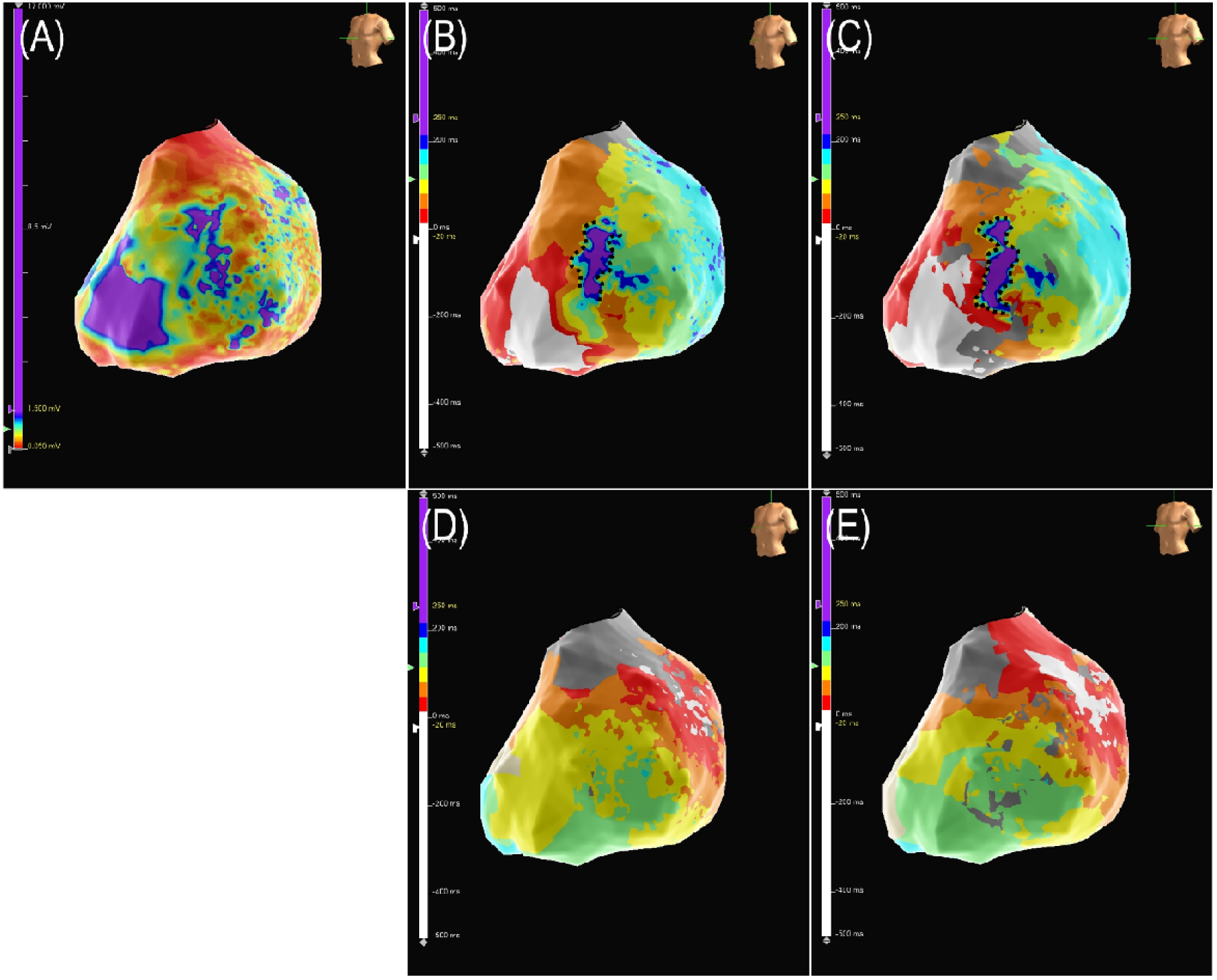
A male in his 70s with nonischemic cardiomyopathy. Epicardial (A) voltage map; (B) RVA S1 map; (C) RVA S2 map; (D) LVOT S1 map; (E) LVOT S2 map. A corridor-shaped LOB (black dotted line) at the apical anterior wall of the LV epicardium was identified in the RVA S1 and RVA S2 maps, but not in the LVOT S1 and LVOT S2 maps. The LOB was not visualized during LVOT pacing because it was parallel to the conduction direction. The LOB was elongated in the RVA S2 map compared with the RVA S1 map. **Abbreviations:** LOB, line of conduction block; LV, left ventricle; LVOT, left ventricular outflow tract; RVA, right ventricular apex.

**Figure 6.**
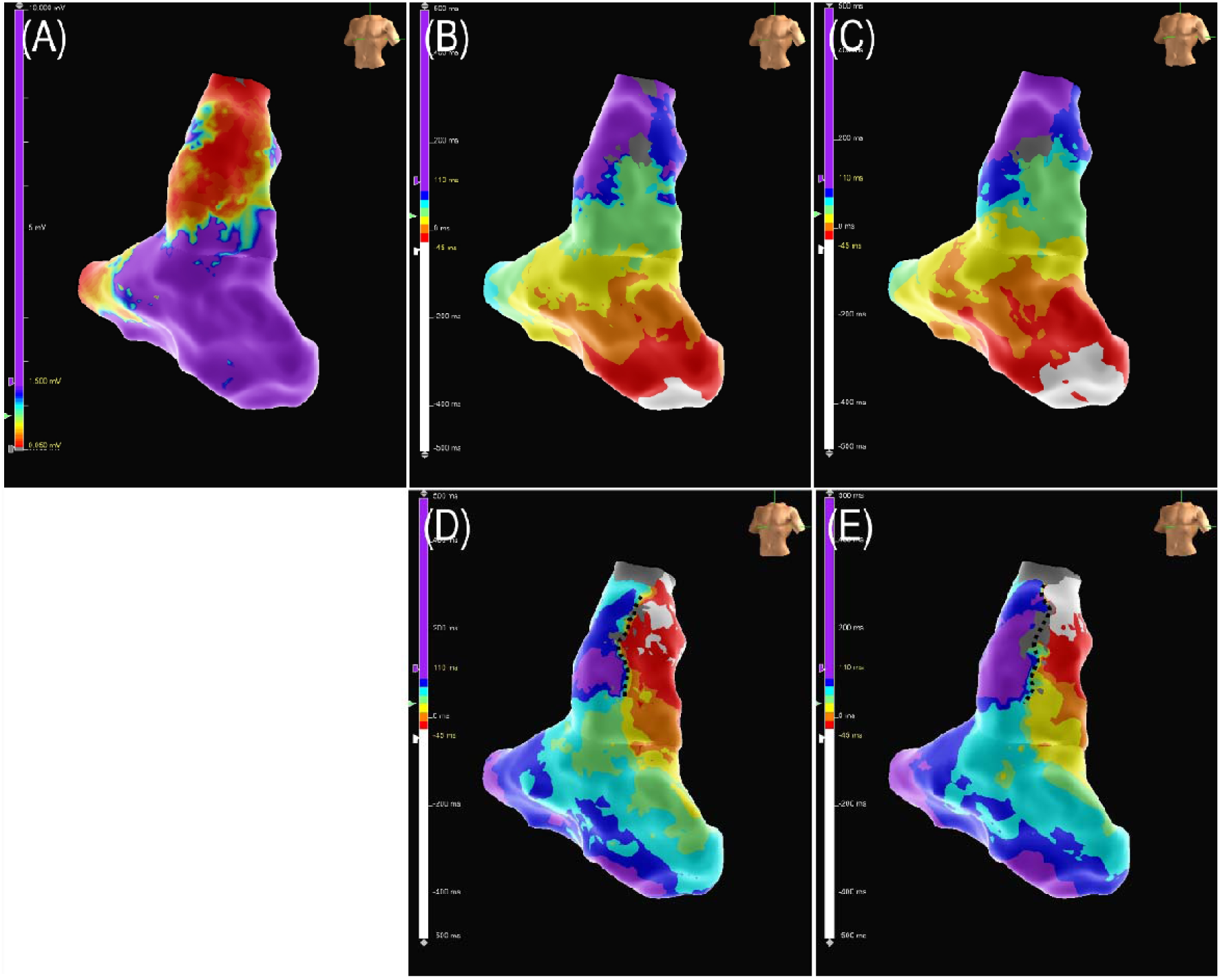
A male in his 50s with nonischemic cardiomyopathy and tetralogy of Fallot. (A) Voltage map; (B) RVA S1 map; (C) RVA S2 map; (D) LVOT S1 map; (E) LVOT S2 map. A LOB (black dotted line) at the RVOT free wall was identified in the LVOT S1 and LVOT S2 maps, but not in the RVA S1 and RVA S2 maps. The LOB was not visualized during RVA pacing because it was parallel to the conduction direction and located at the collision area. **Abbreviations:** LOB, line of conduction block; LVOT, left ventricular outflow tract; RVA, right ventricular apex; RVOT, right ventricular outflow tract.

**Table 4.** Lines of Conduction Block Visible During Only One Pacing Rhythm and Reasons for Invisibility During the Other Pacing Rhythm.

|  | LOB visible only<br>during RVA pacing<br>(n = 2) | LOB visible only<br>during LVOT pacing<br>(n = 6) |
| --- | --- | --- |
| LOB parallel to conduction direction, n (%) | 2 (100.0) | 5 (83.3) |
| LOB at wavefront collision area, n (%) | 0 (0.0) | 1 (16.7) |

**Abbreviations:** LOB, line of conduction block; LVOT, left ventricular outflow tract; RVA, right ventricular apex.

Endpoints were achieved in 78.9% of patients overall, including 80.0% of patients with ICM and 77.8% of patients with NICM. Cardiac tamponade developed in one patient with NICM and was resolved with pericardiocentesis. During a follow-up period of 7.6 ± 3.9 months, VT recurrence occurred in 26.3% of patients—30.0% of those with ICM and 22.2% of those with NICM (Table 2).

## DISCUSSION

### Main Findings

The main findings of the present study are as follows: (1) more LOBs can be identified from the S2 map than from the S1 map; (2) more LOBs can be identified from substrate mapping during LVOT pacing than from RVA pacing; and (3) LOBs located in the wavefront collision area or parallel to the conduction direction may be invisible.

### VT Substrate Mapping

Several substrate mapping methods have been developed. For ILAM, activation mapping is performed during baseline rhythm (sinus rhythm or atrial pacing) by annotating the offsets of ventricular electrograms, and deceleration zones with LOB are identified from the isochronal map.^1^ It has been reported that deceleration zones with LOBs that pacemap match the VT morphology are well correlated with the VT critical isthmus.^8^ LOBs with rotational activation patterns in VT substrate maps are considered more specific findings.^9^

Scar or scar-border tissue with decremental conduction properties may represent potential slow conduction zones within the VT circuit. These decremental conduction areas can become more prominent using the DEEP mapping pacing protocol.^2^ It has been reported that DEEP areas correlate with the VT critical isthmus.^10^ Cardiac MRI scar maps may also be useful for visualizing conduction channels within scar tissue that may represent critical isthmus of VT reentry circuits.^11^ These conduction channels have been shown to correlate well with the VT critical isthmus.^12^

Mapping scar-related VT is more challenging in NICM than in ICM using substrate mapping methods.^13^ This may be because scar tissue in NICM is more heterogeneously or diffusely distributed throughout the ventricle, unlikely the more localized scarring observed in ICM.

### Which Rhythm is Optimal for VT Substrate Mapping?

VT substrate mapping can be performed during sinus rhythm, atrial pacing, RV pacing, LV pacing, and biventricular pacing. The rhythm during mapping can influence the substrate map because the conduction starting point, wavefront direction, and wavefront collision area vary depending on the rhythm. It has been reported that the effectiveness of substrate mapping improves with atrial, RV, or LV pacing.^14^ A pacing rhythm with a pacing site near the scar may be optimal for identifying LOBs.^15^ Sinus rhythm and atrial pacing may not to be optimal for substrate mapping. Because sinus rhythm impulses propagate rapidly along the normal conduction system, the probability of revealing abnormally slow conduction in the LV is relatively low. When the scar zone is activated late, wavefronts may collide at the scar area, rendering LOBs in the wavefront collision area elusive.^15^

Two-site pacing for VT substrate mapping is a reasonable approach because information about the scar and border zones is often insufficient before mapping, and the shapes, directions, and locations of LOBs vary. In addition, several LOBs are three-dimensional hyperboloids.^16^ However, mapping is typically limited to two-dimensional endocardial or epicardial surfaces. We extrapolated the entire structure of the LOBs from the endocardial substrate map, with or without additional epicardial substrate mapping. Two-site pacing substrate mapping may be helpful for extrapolating the entire structure of LOBs.

### DEEP Mapping During RVA and LVOT Pacing

DEEP mapping visualizes more LOBs by evoking decremental conduction using single or double ventricular extrastimuli.^2,17^ In the present study, two-site pacing revealed elusive LOBs by altering wavefront direction and activation timing. However, several challenges exist with two-site pacing DEEP mapping: (1) the mapping process is time-consuming. (2) VT may be consistently induced by DEEP mapping pacing protocol. (3) LOBs unrelated to the VT reentry circuit may be revealed.

To reduce mapping time, substrate mapping during LVOT pacing can be performed first, as more LOBs were identified during LVOT pacing than during RVA pacing in the present study. Based on the voltage map obtained during LVOT pacing, substrate mapping of limited areas (scar and border zones) can subsequently be performed during RVA pacing. To avoid VT storm during substrate mapping, the S2 interval can be extended by 20–30 ms beyond the VT-inducing interval. If VT is easily induced, substrate mapping can be performed during constant-interval pacing.

Pacemapping is essential to exclude LOBs that unrelated to VT circuit. Although there is universally accepted QRS matching threshold for pacemapping, a 90% cutoff was applied in the present study. Because the reentry circuits may partially change or the reentry direction may reverse even when involving the same critical isthmus—setting the cutoff too high may not be optimal. Pacemapping using cycle length of the clinical VT is reasonable because of TR fusion phenomenon.^18^ After substrate mapping and LOB ablation, repeated substrate mapping is important for evaluating ablation effects. It is not uncommon for VT to become hemodynamically stable after substrate ablation.

The present study demonstrated the effects of S2 mapping compared to S1 mapping and highlighted the impact of rhythm changes on substrate mapping. Overall, the findings support the clinical utility of two-site pacing DEEP mapping.

### Study Limitations

This was a single-arm cohort study with a small sample size. Statistical comparison between the ICM and NICM groups were not performed, as this was not the primary aim of the present study. DEEP mapping during both RVA and LVOT pacing was not performed in any of the patients, as hemodynamically unstable VT was easily induced by single or double ventricular extrastimuli in three patients. VT activation mapping was not performed in all patients because VT was hemodynamically unstable in 12 patients, and percutaneous mechanical circulatory support was not available. Therefore, we could not determine whether the LOBs identified from the substrate map corresponded to the critical isthmus of the VT reentry circuits from the VT activation map. However, we confirmed pacemap matching. In addition, the follow-up period was short, and the present study was not designed to evaluate long-term outcomes of VT ablation using two-site pacing DEEP mapping. Large-scale prospective randomized studies on two-site pacing DEEP mapping are warranted.

## Data Availability

The data are available in response to the appropriate requirement.

## Conclusion

A higher number of LOBs could be identified using two-site-pacing (RVA and LVOT) DEEP mapping.

## Acknowledgments

No

## Non-standard Abbreviations and Acronyms

CIED: cardiac implantable electronic device
CRT-D: cardiac resynchronization therapy-defibrillator
DEEP: decrement-evoked potential
HF: heart failure
ICM: ischemic cardiomyopathy
LOB: line of conduction block
LVOT: left ventricular outflow tract
LVEF: left ventricular ejection fraction
NICM: nonischemic cardiomyopathy
RVA: right ventricular apex
RVOT: right ventricular outflow tract
VT: ventricular tachycardia

